# Outpatient COVID-19 convalescent plasma recipient antibody thresholds correlated to reduced hospitalizations within a randomized trial

**DOI:** 10.1101/2023.04.13.23288353

**Authors:** Han-Sol Park, Anna Yin, Caelan Barranta, John S. Lee, Christopher A. Caputo, Jaiprasath Sachithanandham, Maggie Li, Steve Yoon, Ioannis Sitaras, Anne Jedlicka, Yolanda Eby, Malathi Ram, Reinaldo E. Fernandez, Owen R. Baker, Aarthi G. Shenoy, Giselle S. Mosnaim, Yuriko Fukuta, Bela Patel, Sonya L. Heath, Adam C. Levine, Barry R. Meisenberg, Emily S. Spivak, Shweta Anjan, Moises A. Huaman, Janis E. Blair, Judith S. Currier, James H. Paxton, Jonathan M. Gerber, Joann R. Petrini, Patrick B. Broderick, William Rausch, Marie Elena Cordisco, Jean Hammel, Benjamin Greenblatt, Valerie C. Cluzet, Daniel Cruser, Kevin Oei, Matthew Abinante, Laura L. Hammitt, Catherine G. Sutcliffe, Donald N. Forthal, Martin S. Zand, Edward R. Cachay, Jay S. Raval, Seble G. Kassaye, Christi E. Marshall, Anusha Yarava, Karen Lane, Nichol A. McBee, Amy L. Gawad, Nicky Karlen, Atika Singh, Daniel E. Ford, Douglas A. Jabs, Lawrence J. Appel, David M. Shade, Bryan Lau, Stephan Ehrhardt, Sheriza N. Baksh, Janna R. Shapiro, Jiangda Ou, Yu Bin Na, Maria D. Knoll, Elysse Ornelas-Gatdula, Netzahualcoyotl Arroyo-Curras, Thomas J. Gniadek, Patrizio Caturegli, Jinke Wu, Nelson Ndahiro, Michael J. Betenbaugh, Alyssa Ziman, Daniel F. Hanley, Arturo Casadevall, Shmuel Shoham, Evan M. Bloch, Kelly A. Gebo, Aaron A.R. Tobian, Oliver Laeyendecker, Andrew Pekosz, Sabra L. Klein, David J. Sullivan

## Abstract

**BACKGROUND:** The COVID-19 convalescent plasma (CCP) viral specific antibody levels that translate into recipient post-transfusion antibody levels sufficient to prevent disease progression is not defined.

**METHODS:** This secondary analysis correlated donor and recipient antibody levels to hospitalization risk among unvaccinated, seronegative CCP recipients within the outpatient, double blind, randomized clinical trial that compared CCP to control plasma. The majority of COVID-19 CCP arm hospitalizations (15/17, 88%) occurred in this unvaccinated, seronegative subgroup. A functional cutoff to delineate recipient high versus low post-transfusion antibody levels was established by two methods: 1) analyzing virus neutralization-equivalent anti-S-RBD IgG responses in donors or 2) receiver operating characteristic (ROC) analysis.

**RESULTS:** SARS-CoV-2 anti-S-RBD IgG antibody was diluted by a factor of 21.3 into post-transfusion seronegative recipients from matched donor units. Viral specific antibody delivered approximated 1.2 mg. The high antibody recipients transfused early (symptom onset within 5 days) had no hospitalizations. A CCP recipient analysis for antibody thresholds correlated to reduced hospitalizations found a significant association with Fisher’s exact test between early and high antibodies versus all other CCP recipients (or control plasma) with antibody cutoffs established by both methods-donor virus neutralization-based cutoff: (0/85; 0% versus 15/276; 5.6%) p=0.03 or ROC based cutoff: (0/94; 0% versus 15/267; 5.4%) p=0.01.

**CONCLUSION:** In unvaccinated, seronegative CCP recipients, early transfusion of plasma units corresponding to the upper 30% of all study donors reduced outpatient hospitalizations. These high antibody level plasma units, given early, should be reserved for therapeutic use.

Trial registration: NCT04373460

**FUNDING:** Defense Health Agency and others.

## INTRODUCTION

The SARS-CoV-2 specific antibody levels necessary to prevent infection or reduce hospitalization from mild outpatient COVID-19 or reduce deaths in those already hospitalized are likely to be different. For hospitalized patients, effective CCP antibody levels have been estimated from registries(1, 2), but comparable information is not available for outpatient usage. The high inter-laboratory variability with diverse SARS-CoV-2 serologic assays for binding or virus neutralization antibody levels creates further challenges(3, 4). Dilutional live or pseudovirus neutralization measures from 27 separate pre-alpha convalescent plasma collections varied in geometric means for 50% inhibition from 19 to 4,344 with a mean of 311(5). Separating protective antibody metrics in vaccinated people or COVID-19 convalescent plasma donors that are still therapeutic after dilution further adds to complexity. For example, influenza vaccinees in the 1970s with dilutional virus hemagglutination inhibition titer ≥1:40 prevented infection(6, 7), such that the World Health Organization set the threshold of protection at 1:40(8). Infants with Respiratory Syncytial Virus in two separate studies with neutralizing antibody titers over 1:256 are protected from hospitalizations(9, 10). However, therapeutic convalescent plasma would need to have 10-20 times the protective neutralization titer after a small plasma volume is diluted into a seronegative recipient.

We previously reported that outpatient transfusion randomized to CCP or control plasma in 1181 participants with pre-delta CCP reduced the risk of hospitalization by 54%(11). A prespecified analysis from the parent outpatient CCP RCT aimed to compare antibody levels in donor-recipient pairs to explore the association between antibody levels and prevention of hospitalizations in recipients. With 88% of post-transfusion COVID-19 hospitalizations (15 of 17 total) occurring among unvaccinated, seronegative outpatient recipients, we analyzed hospitalization risk among this group by comparing CCP recipients stratified by early or late treatment (i.e., ≤ 5 versus >5 days from symptom onset) with antibody levels to demarcate pre-delta CCP for pre-omicron recipient thresholds for efficacy to reduce mild CoVID-19 hospitalizations.

## RESULTS

### Trial population

This secondary analysis includes the unvaccinated at screening subgroup to correlate donor and post-transfusion antibody levels with hospitalization. Transfusions spanned 16 months from June 3, 2020 to Oct 1, 2021 with last three month follow up after transfusion in January 2022.

The unvaccinated seropositive rate was 21%. Among the seronegative, unvaccinated patients analyzed, 368 received control plasma and 366 received CCP with an average age of 44 years old. Both control and CCP arms were predominately female, obese (44% with BMI ≥ 30), and had at least one pre-existing comorbidity (41%). All COVID-19-related hospitalizations in the CCP arm recipients (17 total) were among unvaccinated recipients—15 seronegative (88%) and 2 seropositive recipients (12%; **Figure 1**, **Table 1**). Excluded from this analysis were the 159 fully vaccinated with no hospitalizations, 58 partly vaccinated with one hospitalization and 199 unvaccinated seropositive with 7 hospitalizations.

**Figure 1.**
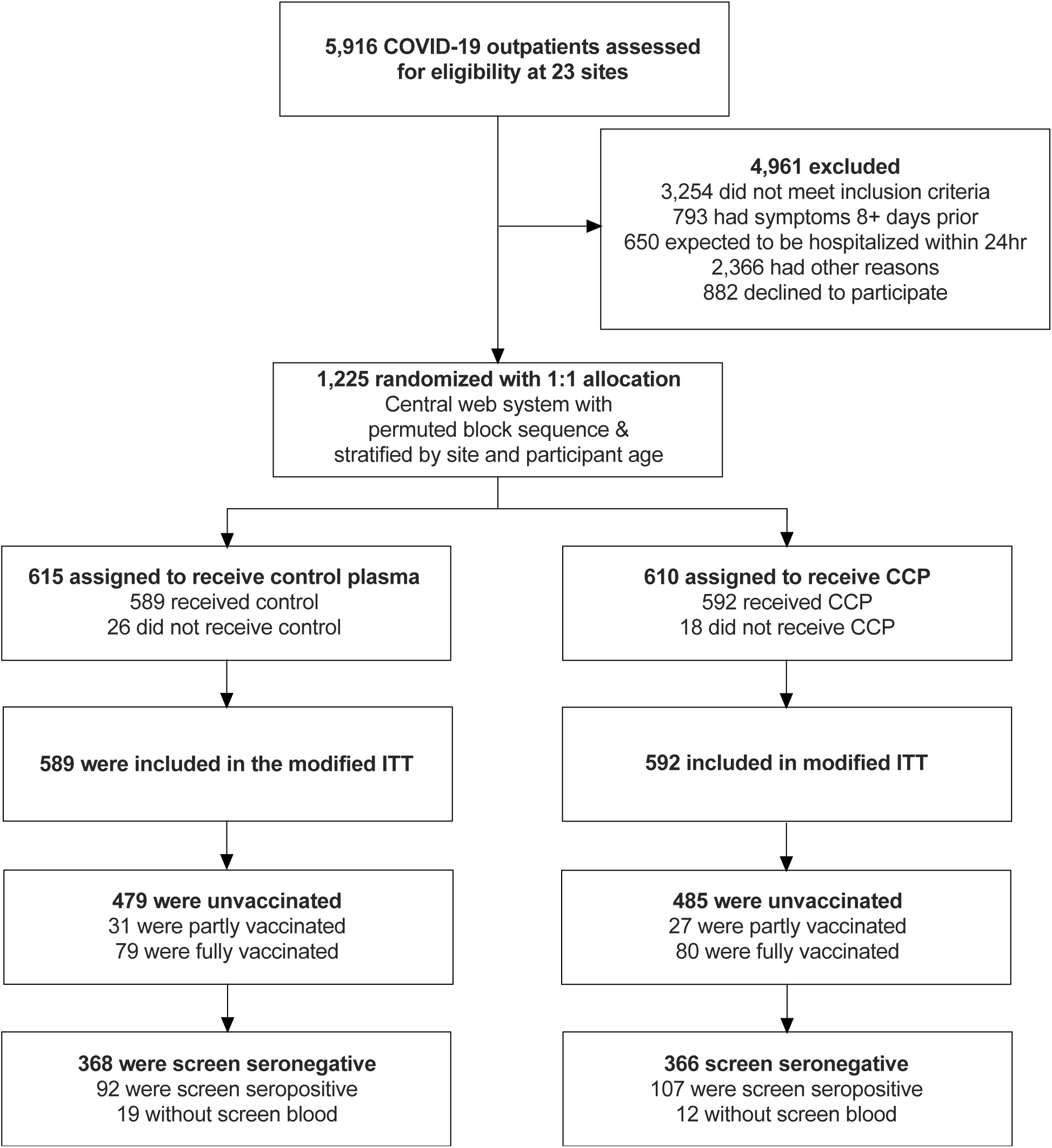
CONSORT diagram depicting enrollment, allocation, and analysis flow of recipients.

**Table 1.** Unvaccinated control and convalescent plasma recipient demographics and comorbidities.

|  | **Total** | **Control** | **CCP** | **Control**<br>**Early** | **Control**<br>**Late** | **CCP**<br>**Early** | **CCP**<br>**Late** |
| --- | --- | --- | --- | --- | --- | --- | --- |
| **Number** | 964 | 479 | 485 | 207 | 272 | 210 | 275 |
| **Age, mean (SD)** | 44 (14) | 45 (14) | 43 (14) | 45 (14) | 45 (14) | 43 (14) | 44 (15) |
| **Sex, n (%)** |  |  |  |  |  |  |  |
| Female | 545 (57) | 283 (59) | 262 (54) | 115 (56) | 168 (62) | 113 (54) | 149 (54) |
| Male | 419 (43) | 196 (41) | 223 (46) | 92 (44) | 104 (38) | 97 (46) | 126 (46) |
| **Race, n (%)** |  |  |  |  |  |  |  |
| Asian | 33 (3) | 16 (3) | 17 (4) | 12 (6) | 4 (1) | 4 (2) | 13 (5) |
| Black | 136 (14) | 63 (13) | 73 (15) | 27 (13) | 36 (13) | 26 (12) | 47 (17) |
| American Indian | 16 (2) | 8 (2) | 8 (2) | 6 (3) | 2 (1) | 3 (1) | 5 (2) |
| Pacific Islander | 4 (0) | 2 (0) | 2 (0) | 1 (0) | 1 (0) | 0 (0) | 2 (1) |
| Other race/not reported | 4 (0) | 7 (1) | 9 (2) | 4 (2) | 3 (1) | 6 (3) | 3 (1) |
| White | 759 (79) | 383 (80) | 376 (78) | 157 (76) | 226 (83) | 171 (81) | 205 (75) |
| **Ethnicity, n (%)** |  |  |  |  |  |  |  |
| Hispanic or Latino | 143 (15) | 72 (15) | 71 (15) | 37 (18) | 35 (13) | 33 (16) | 38 (14) |
| **BMI Category, n (%)** |  |  |  |  |  |  |  |
| BMI <30 | 542 (56) | 261 (54) | 281 (58) | 112 (54) | 149 (55) | 122 (58) | 159 (58) |
| BMI ≥30 | 422 (44) | 218 (46) | 204 (42) | 95 (46) | 123 (45) | 88 (42) | 116 (42) |
| **Hypertension, n (%)** | 242 (25) | 117 (24) | 125 (26) | 52 (25) | 65 (24) | 55 (26) | 70 (25) |
| **Diabetes, n (%)** | 85 (9) | 47 (10) | 38 (8) | 19 (9) | 28 (10) | 13 (6) | 25 (9) |
| **Asthma, n (%)** | 110 (11) | 59 (12) | 51 (11) | 25 (12) | 34 (13) | 24 (11) | 27 (10) |
| **HIV, n (%)** | 23 (2) | 12 (3) | 11 (2) | 4 (2) | 8 (3) | 6 (3) | 5 (2) |
| **Pregnancy, n (%)** | 2 (0) | 0 (0) | 2 (0) | 0 (0) | 0 (0) | 1 (0) | 1 (0) |
| **Hospitalizations, n (%)** |  |  |  |  |  |  |  |
| Total unvaccinated | 53 (6) | 36 (8) | 17 (4) | 24 (12) | 12 (4) | 5 (2) | 12 (4) |
| Seronegative | 46 (5) | 31 (6) | 15 (3) | 20 (10) | 11 (4) | 5 (2) | 10 (4) |
| Seropositive | 7 (1) | 5 (1) | 2 (0) | 4 (2) | 1 (0) | 0 (0) | 2 (0) |
| **Serostatus at Screen, n (%)** |  |  |  |  |  |  |  |
| Seronegative | 734 (76) | 368 (77) | 366 (75) | 167 (81) | 201 (74) | 173 (82) | 193 (70) |
| Seropositive | 199 (21) | 92 (19) | 107 (22) | 30 (14) | 62 (23) | 34 (16) | 73 (27) |
| No screen bloods | 31 (3) | 19 (4) | 12 (2) | 10 (5) | 9 (3) | 3 (1) | 9 (3) |
| **Viral Copies, GM** | 8,718 | 7,389 | 10,245 | 17,660 | 3,817 | 35,698 | 3,834 |
| Hospitalized | 6,346 | 4,770 | 10,501 | 7,114 | 2,145 | 65,686 | 4,892 |
| Non-Hospitalized | 8,872 | 7,627 | 10,235 | 19,641 | 3,909 | 35,144 | 3,788 |

### CCP donor antibody levels

Approximately 40% of all potential CCP research study donors in the parent study were excluded due to low antibody levels. Unique transfusion units represented the upper 60% of all CCP pre-delta donors with a geometric mean (GM) anti-S-RBD IgG 1:6,741 titer (3,161 AUC).

Donor plasma showed strong correlations between anti-S-RBD IgG and virus neutralization antibody (nAb) in dilutional titer and AUC (**Figure 2A**), as well as donor virus-specific anti-S-RBD IgG antibodies in ng/mL with anti-S-RBD IgG AUC (**Supplemental Figure 1**). We estimate that the total viral-specific anti-S-RBD IgG dose from donor into recipient is 1.2mg based on a transfusion volume of 200mL with a donor anti-S-RBD IgG GM of 5.1 μg/mL, indicating recipients have low post-transfusion antibody levels based on current dosing recommendation for CCP (**Supplemental Figure 1**, **Supplemental Table 1**).

**Figure 2.**
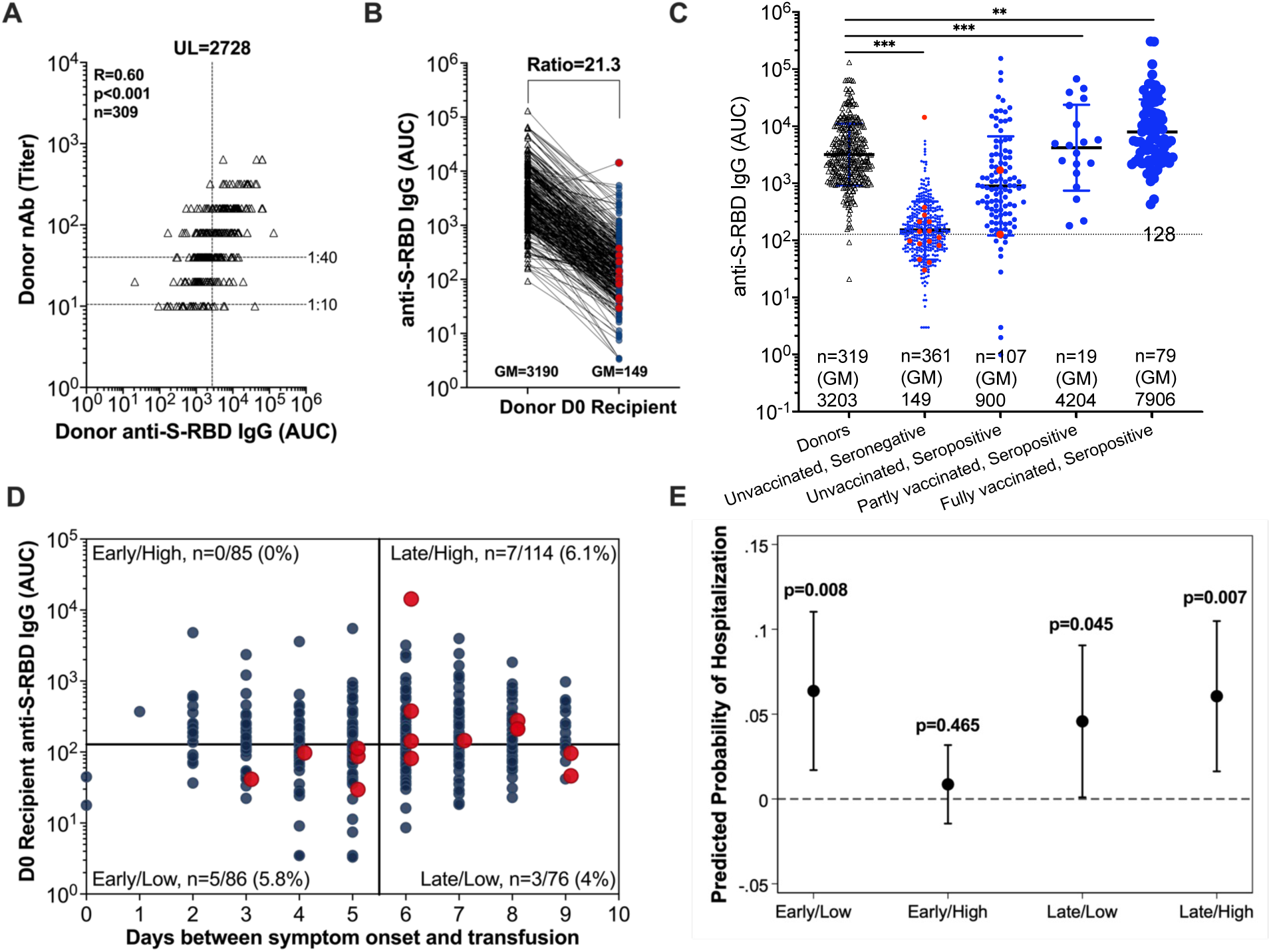
Utilizing CCP donor neutralizing antibody and anti-S-RBD levels to establish a functional cutoff associated with hospitalization protection among their respective screened seronegative recipients. (A) A 1:40 dilutional titer for virus neutralizing antibody (nAb) was previously identified as a correlate of protection in influenza studies. Here, we use the 1:40 dilutional titer for nAb to identify the upper limit level of donor anti-S-RBD IgG 2728 AUC associated with protection from hospitalization. 1:10 dilutional titer is the limit of detection for nAb. (B) Ratio of matched donor anti-S-RBD IgG AUC to that of their respective CCP seronegative recipients that was used to infer the functional cutoff in recipients was determined to be 21.3 (C) Anti-S-RBD IgG AUC levels among donors and post-transfusion recipients segregated by screen vaccination status and serostatus compared by Kruskall-Wallis with Dunn’s post-hoc corrections, *p<0.033, **p<0.002, ***p<0.001. Unvaccinated subsequently hospitalized (red dots) post transfusion recipients in screen seronegative (n=15) and screen seropositive (n=2). (D) Screen seronegative, unvaccinated recipient D0 (post-transfusion) antibody (n=361) segregated by recipient days from symptom onset to transfusion and high (>128 AUC) or low (≤ 128 AUC) levels. Recipient high and low cutoffs were calculated by using a 21.3-fold drop from the donor cutoff determined by the upper value of the 95% confidence interval for the geometric mean of donor anti-S-RBD AUC at a 1:40 nAb titer associated with protection. Subsequently hospitalized (red dot) and non-hospitalized (blue dot) recipients are shown. (E) Firth logistic regression model adjusted for age, sex, BMI, and variant to compare the predicted probabilities of hospitalization across early vs. late and high (>128 AUC) vs. low (≤ 128 AUC) categories of screen seronegative, unvaccinated CCP recipients. The horizontal dashed line represents a predicted probability of 0%. P-values for the predicted probability of each category are shown with p<0.05 considered significant.

### Screen and post-transfusion antibody levels among unvaccinated, seronegative recipients

The dilution factor associated with the administration of approximately 200mL CCP was determined by comparing the GM of anti-S-RBD IgG AUC levels of matched donors to that of unvaccinated seronegative recipients. The donor anti-S-RBD IgG AUC GM (3,190) proportionately decreased by a factor of 21.3 when compared to the anti-S-RBD IgG AUC GM (149) for seronegative recipients AUC within 30 minutes of transfusion (**Figure 2B**). Similarly, 15 seronegative hospitalized CCP recipients had post-transfusion antibody levels 19 times lower than matched donors. The hospitalized and non-hospitalized unvaccinated, screened seropositive CCP participants had a post-transfusion GM anti-S-RBD-IgG 836 AUC, with those partly vaccinated at 4,204 AUC and those fully vaccinated breakthrough infection at 7,908 AUC (**Figure 2C**). The pre-transfusion antibody levels of unvaccinated seropositive participants increased with the days from symptom onset to transfusion (**Supplemental Figure 2**).

### Post-transfusion recipient antibody benchmarks associated with no hospitalization

Among seronegative control recipients, 7.6% (28/368) were hospitalized, which was higher than the 6.3% hospitalization rate among controls of the parent study that included vaccinated (full and partial) and nonvaccinated, seropositive participants. Hospitalizations in all seronegative CCP recipients were 4.1% (15/366), slightly higher than the full study finding of 2.9%.

For this subgroup analysis, we estimated the antibody threshold levels correlated to protection from hospital progression in the CCP group for early and late transfusions—one based on binding antibody levels associated with functional virus neutralization (**Figure 2**) and another by ROC analysis (**Figure 3**). For the functional cutoff based on virus nAb, we used a 40-fold dilution of virus nAb, like the correlate of infection protection previously reported for influenza(8). By plotting donor anti-S-RBD-IgG AUC against increasing 2-fold viral dilutions, we identified donor anti-S-RBD IgG 2,728 AUC as the upper limit of the 95% confidence interval of the GM at a 40-fold nAb titer (**Figure 2A**). After a 21.3 dilution, the post-transfusion threshold was calculated to be 128 AUC in recipients. These functional cutoffs delineate high versus low anti-S-RBD IgG levels at 2,728 and 128 AUC for donors and their matched unvaccinated, seronegative recipients, respectively. Recipient post-transfusion antibody levels were plotted by days between symptom onset to transfusion to correlate the functional cutoffs with hospitalization outcome (**Figure 2D-E**).

**Figure 3.**
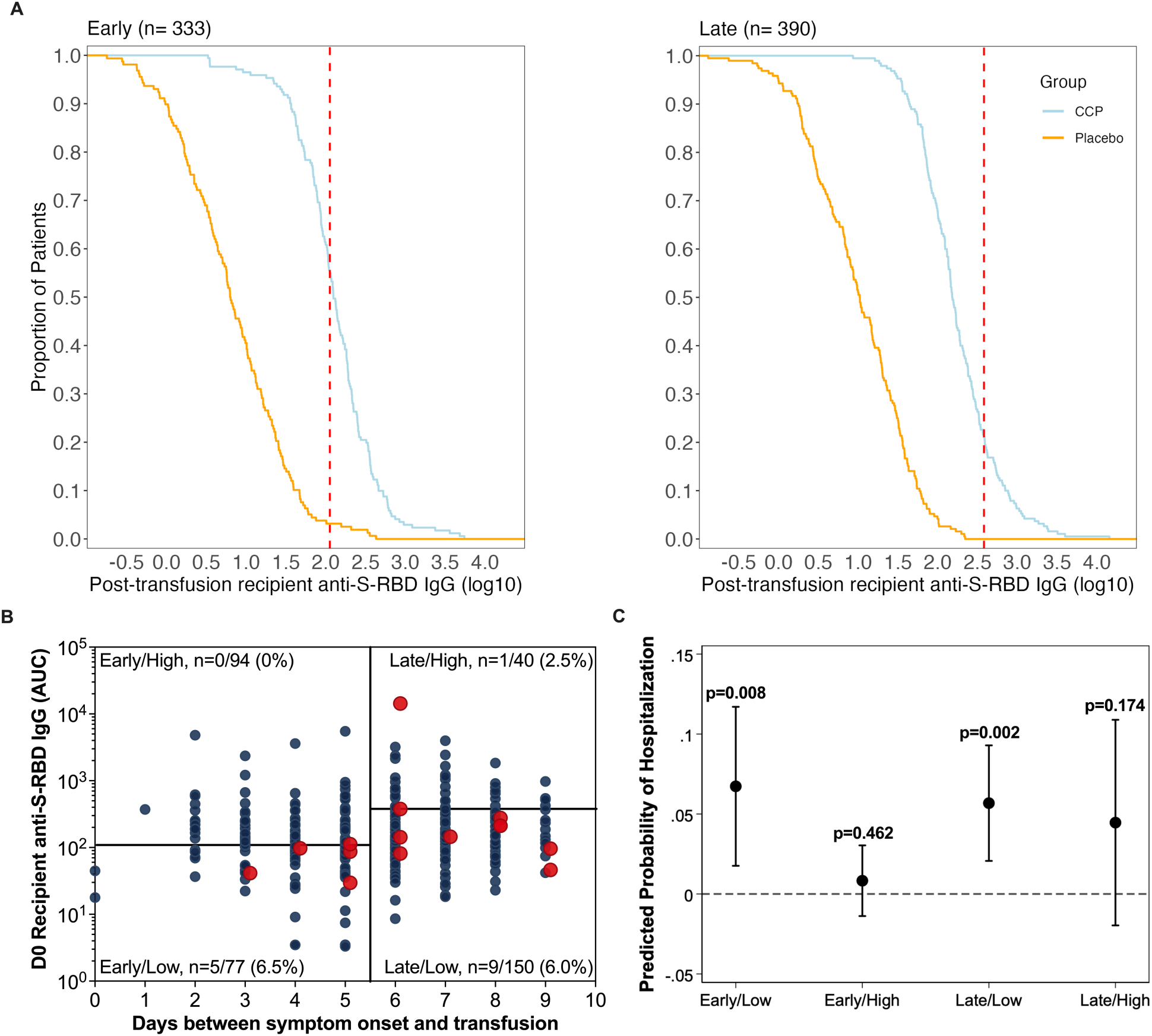
CCP recipient D0 post-transfusion and matched donor antibody levels stratified by duration from symptom onset to transfusion using cutoffs established by the ROC and maximum antibody threshold method. (A) RCDC curves were used to compare the maximum antibody thresholds, as represented by the red dashed lines, to delineate high and low antibody levels for early or late CCP recipients. Early recipients are delineated at antilog of 2.06 anti-S-RBD AUC (115 AUC) while late recipients are delineated at antilog of 2.58 anti-S-RBD AUC (380.2 AUC). Curves exclude five early and one late control participant(s) whose post-transfusion plasma was not available. (B) Screen seronegative, unvaccinated recipient D0 post-transfusion antibody (n=361) segregated by recipient days from symptom onset to transfusion based on high versus low cutoffs established by the ROC and maximum antibody threshold method. Subsequently hospitalized (red) and non-hospitalized (blue) recipients are shown. (C) Firth logistic regression model adjusted for age, sex, BMI, and variant to compare the predicted probabilities of hospitalization across early vs. late and high vs. low categories of screen seronegative, unvaccinated CCP recipients based on the ROC and maximum antibody threshold method. The horizontal dashed line represents a predicted probability of 0%. P-values for the predicted probability of each category are shown with p<0.05 considered significant.

### Virus neutralization-based correlate of protection from hospitalization in recipients

We observed zero hospitalizations among recipients transfused early (i.e., ≤ 5 days after symptom onset) with post-transfusion anti-S-RBD IgG levels above 128 AUC as compared to the other three CCP quadrants. Although the probability of hospitalization was lowest among recipients receiving early transfusion and high antibody levels above 128 AUC, this group did not reach statistical significance when compared to the other quadrants by Firth’s logistic regression, potentially due to smaller sample sizes (**Figure 2E; Supplemental Table 2**).

Exploratory analysis with Fisher’s exact test revealed a significant association between early/high transfusion, as defined by the nAb-based method, with hospitalization status among other unvaccinated, seronegative CCP recipients (p=0.03), indicating a difference in probability of hospitalization between those with early/high CCP transfusion (0/85, 0%) and those early/low or late CCP (15/276, 5.6%). The early/high CCP compared to all controls (28/368, 7.6%;p=0.004) or early controls (18/167, 11.7%;p=0.0005) was even more significant(**Supplemental Table 3)**.

### ROC-based correlate of protection from hospitalization in recipients

As an alternative method for identifying antibody thresholds for early recipients, ROC analysis with maximum percent hospital reduction were used to determine the antibody threshold level for late transfusions. The red dotted line in RCDC demarcates early transfusion ROC 115 anti-S-RBD IgG AUC maximum while the late transfusion 380 AUC maximized hospitalization difference (**Figure 3A**). Hospitalization was reduced (0 of 94 hospitalized) with anti-S-RBD 115 AUC (log10 of 2.06), while for recipients treated after 5 days from symptom onset, the antibody level for similar treatment efficacy (1 of 40) was 380 AUC (log10 of 2.58; **Figure 3B**). A Firth’s logistic regression comparing CCP quadrants revealed that recipients receiving early transfusion with high post-transfusion antibody levels above 115 AUC had the lowest probability of hospitalization, but this difference from other quadrants was not statistically significant (**Figure 3C**). Exploratory analysis with Fisher’s exact test revealed a significant association between early/high transfusion, as defined by the RCDC-based method, with hospitalization status among unvaccinated, seronegative CCP recipients (p=0.01), indicating a difference in probability of hospitalization between those with early/high transfusion (0/94, 0%) and those early/low or late CCP (15/267, 5.4%). The ROC early/high CCP compared to all controls (28/368, 7.6%;p=0.002) or early controls (18/167, 11.7%;p=0.0005) had greater significance **(Supplemental Table 3)**.

### Donor antibody-based correlate of protection from hospitalization

The early/high quadrant for donor plasma units based on the 2768 AUC (1/88-1.1%) was also found to be significantly different by Fishers exact test from all seronegative controls (28/368-12%)(p<0.002) and early seronegative controls(18/167-10.7%) (p<0.002; **Supplemental Table 3**). Donor plasma antibody-based relative risk reduction is 86% and absolute risk reduction is 6.5%. Comparison of donor early/high units to early/low and both late CCP was not significant (**Supplemental Table 3**).

### Nasal SARS-CoV-2 viral RNA copies at screening

Nasal viral load might independently determine risk of hospitalization. All unvaccinated individuals subsequently receiving either control plasma or CCP had indistinguishable screen (before plasma transfusion) nasal viral loads, regardless of subsequent hospitalization outcome (**Table 1, Supplemental Figure 3A**). Nasal viral loads of those receiving early transfusions were associated with higher viral loads compared to late transfusions, regardless of serostatus at screening (**Supplemental Figure 3B-C**). Delta period viral loads in unvaccinated and vaccinated individuals showed a similar drop in viral load with later transfusions (**Supplemental Figure 4**). While our inclusion criteria required a documented positive molecular SARS-CoV-2 test (87% by RNA detection and 13% by antigen detection), the interval between subjects’ initial pre-enrollment SARS-CoV-2 test and our pre-transfusion nasal swab may have been up to 7 days.

### Longitudinal antibody kinetics following transfusion

Antibody levels at or beyond 14 days post-transfusion did not differ between CCP and control plasma recipients (**Figure 4**). Hospitalization status, but not treatment, affected antibody levels over time. The multivariable linear mixed-effects regression, adjusted for variant, age, sex, and BMI, showed no differences in antibody levels between CCP and control plasma recipients beyond 14 days post-transfusion (**Figure 4**). There were neither sex, age, BMI, nor comorbidity differences in antibody levels between CCP and control groups. At the day 90 follow-up visit, anti-S-RBD IgG AUC levels were similar for control and CCP and increased during variant periods pre-alpha, alpha and delta as well as among fully vaccinated recipients (**Supplemental Figure 5**).

**Figure 4.**
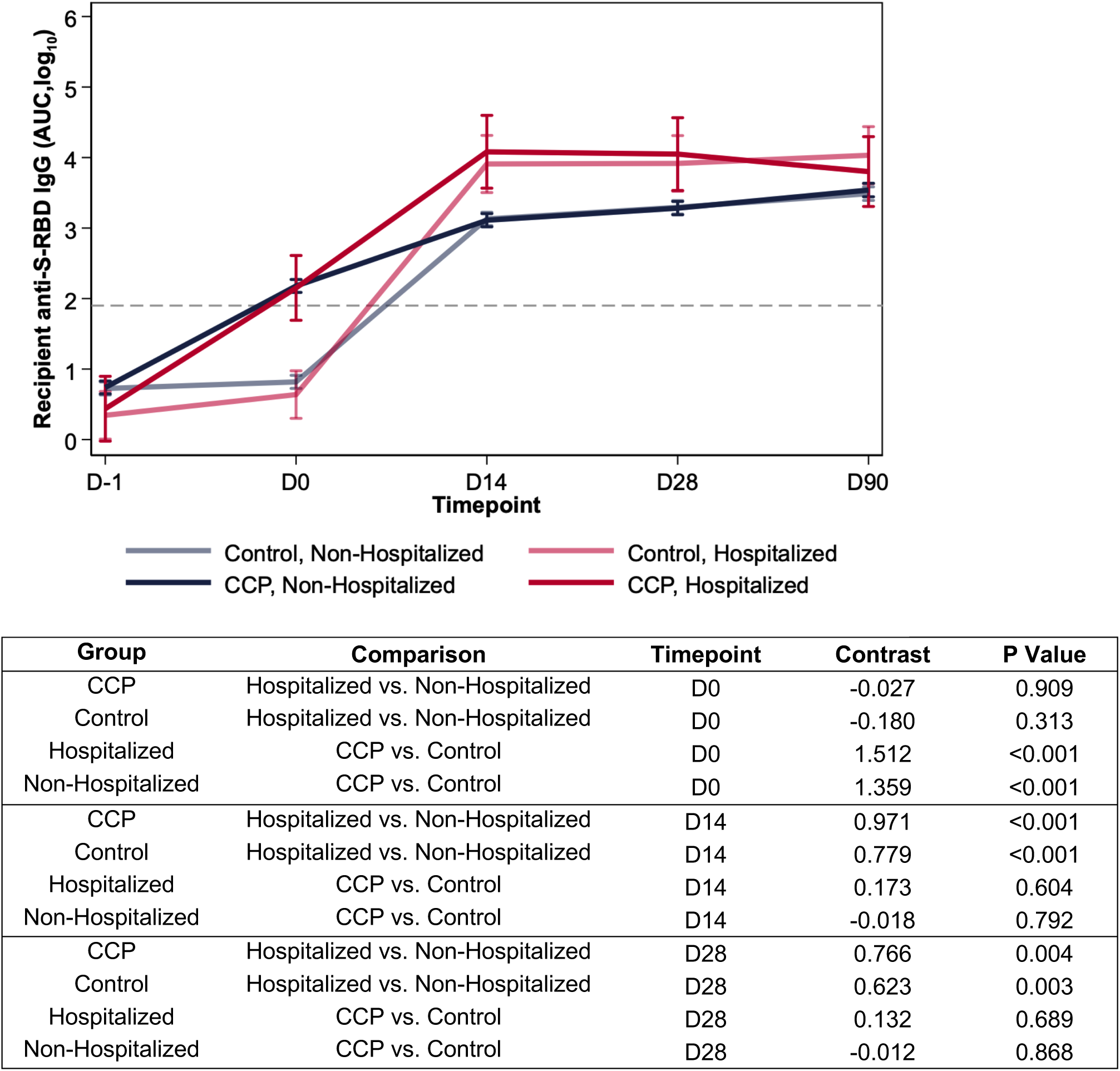
Antibody levels over three months post-transfusion by hospitalization status and treatment group for screen seronegative, unvaccinated recipients. Log_10_-transformed antibody levels up to 90 days post-transfusion were segregated by treatment and hospitalization status of recipients using a linear mixed effects regression model, adjusted for variant, age, sex, and BMI. CCP recipients have greater AUC levels on D0, but by D14, the hospitalized recipients have greater AUC levels than non-hospitalized. The average days from transfusion to hospitalization was 3.05 days, with all post-transfusion hospitalizations occurring between D0 and D14. The dashed line represents the log-transformed cutoff (1.924) for seropositivity. This diagnostic threshold is equivalent to the anti-S-RBD IgG log_10_-transformed 180 titer.

## DISCUSSION

In this secondary analysis of our outpatient, double-blind, placebo-controlled trial of CCP to prevent hospitalizations, we documented that donor CCP in the top 30% by antibody levels increased seronegative recipient antibody thresholds sufficiently to high cut-offs that when administered early within 5 days of symptom onset were effective in hospital reduction. Initial screen nasal viral loads did not impact hospital outcome.

At the start of the pandemic there were no evidence-based standards for CCP donor antibody levels. Most donor EUA qualification of high titer after February 2021 was based on anti-spike antibody levels rather than neutralizations. Diversity in methods used antibody quantification and the need for harmonization of assays across institutions became apparent(12, 13). Within our study, the donor binding anti-S-RBD IgG of 2,728 AUC corresponded to live virus neutralization of greater than 1:40 in donors, and if transfused within 5 days of COVID-19 symptom onset, reduced hospitalization. Initially, the FDA recommended donor plasma qualification for the outpatient CCP study under IND19725 as seropositive after a 1:320 dilution(11). CCP donors for the hospitalized Expanded Access Program from March to August 2020 in the USA reported more than ten-fold higher median virus neutralization, using the Broad Institute Plaque reduction neutralization test (D614G), of 1:525(2). The outpatient C3PO study used microneutralization assay with a median 1:578 titer(14). The Argentina outpatient study used a CCP cutoff of 1:84 based on a surrogate virus neutralization test and segregating to upper half of donors improved outcome(15). The effective CONFIDENT trial used CCP with virus neutralization of greater than 1:160 representing the top 15% of Belgium donors in the pre-delta time periods in those hospitalized and newly mechanically ventilated(16). While the lack of standardization impedes comparative virus neutralizations analysis, all studies highlight that utilizing donors with high titer virus neutralizing antibodies is critical for CCP effectiveness.

When CCP was first deployed in 2020, there were concerns that specific antibody administration to individuals in the early stages of COVID-19 could interfere with the development of endogenous immune responses(17). However, our findings show that transfusion of CCP, as compared to control plasma, was not associated with differences in the total antibody level immune response in recipients, reassuring for the immunological safety of CCP in humans. The C3PO convalescent plasma study also demonstrated no antibody level difference between CCP and saline infusions(14, 18).

Strengths of this study include the large participant population of 1181, well-characterized donor and recipient antibody levels measured by diverse metrics and overall trial effectiveness in hospital reduction that extended to subpopulations at risk of severe disease progression like diabetes, hypertension, obesity and increasing age. Limitations to the study include predominately SARS-CoV-2 naïve recipients enrolled prior to the Omicron variant who were largely unvaccinated such that the findings are only approximately applicable to immunocompromised patients or others who lack SARS-CoV-2 antibodies. Another limitation is the low number of seronegative participants transfused within 5 days of symptom onset with post-transfusion donor antibody levels above the geometric mean in our study population (approximately 100 participants). The study randomized participants to CCP and control plasma, not early or late transfusions stratified by antibody level. The parent study was not powered to look at these stratified quadrants.

Our results provide evidence for the best use of CCP. In summary, our results support and confirm that for antibody therapy to be effective, sufficient levels of pathogen specific antibodies need administration early(19). The mass amount of antibody needed for CCP efficacy (1.2 mg) is much lower than when mAbs were used (150 mg to 2100 mg), which may reflect synergy between the many antibody specificities and isotypes in the polyclonal response. No hospitalizations were observed in those recipients treated within 5 days of symptom onset with high antibody levels, indicating that this is the optimal dose and timing combination for effective CCP use. Early treatment alone is insufficient, as hospitalizations were still observed in the group treated within 5 days with lower titer units, necessitating both early treatment and adequate antibody dosing for optimal efficacy.

Although our results are less relevant to COVID-19 in the fourth year of the pandemic when the majority of immunocompetent individuals have endogenous antibody from vaccination and/or infection, they are highly relevant to both the currently immunocompromised COVID-19 patients without functional SARS-CoV-2 antibodies or to future deployments of convalescent plasma for infectious disease emergencies. We advocate that CCP units reserved for therapy comprise greater antibody levels restricted to the upper 20-30% of all donors to protect against future variants(20–22) or a novel microbe.

When humanity faces its next pandemic, there is a high likelihood that convalescent plasma will be used again until better specific therapies become available. Our data provide a roadmap for optimal early, high dose (upper deciles) convalescent plasma deployment with future emergencies.

## METHODS

### Study Design

This study is a follow-up secondary analysis to correlate donor and recipient antibody levels to hospital outcome within a large outpatient, double-blind, randomized clinical trial comparing CCP to control plasma at 23 centers throughout the United States from June 2020 through September 2021(11). Symptomatic adults (≥18 years old) with a confirmed SARS-CoV-2 positive test, regardless of vaccination status or risk factors for severe COVID-19, were enrolled within 8 days of symptom onset. Over 5,000 recipient plasma samples were collected at pre-transfusion screening (D-1), 30 minutes post-transfusion (D0), and follow-up visits (D14, D28, D90)(23). This subgroup analysis was restricted to seronegative, unvaccinated CCP recipients. CONSORT reporting guidelines were utilized(24). Detailed methods are in the Supplement Appendix.

### Study Donor Plasma

The study qualified donor plasma with SARS-CoV-2 positive antibodies after a 1:320 dilution under FDA IND 19725 protocol. After July 2021, the transfused plasma donor units met the existing FDA Emergency Use Authorization (EUA) criteria for high titer at EUROIMMUN arbitrary unit (AU) over 3.5. These donor units were previously characterized for full-length anti-Spike IgG geometric mean (GM) titers of 13,053, which corresponded with a more precise area under the curve (AUC) geometric mean of 7938, equaling 243 BAU/mL using the international standards(25). The median neutralizing antibody (nAb) titer was 80, with a geometric mean titer of 58, and nAb AUC of 51, equaling GM 27 IU/mL(25). The commercial EUROIMMUN arbitrary units (AU) mean was 6 for the unique donor units(25).

### Study Approval

Johns Hopkins University (JHU) served as the single-IRB (sIRB). For the Center for American Indian Health sites, the protocol was also independently reviewed and approved by the Navajo Nation Health Human Research Review Board and the Indian Health Service IRB. The protocol was also approved by the Department of Defense (DoD) Human Research Protection Office (HRPO). The trial was conducted in accordance with the principles of the Declaration of Helsinki, the Good Clinical Practice guidelines of the International Council for Harmonization, and all applicable regulatory requirements. Written and signed informed consent were obtained from all participants. The trial was registered on clinical trials.gov- NCT04373460.

### Indirect anti-S-RBD ELISA

The anti-S-RBD ELISA was adapted from a published protocol(26). The anti-S-RBD IgG threshold for seronegativity was 180 titer or below. Serostatus was determined based on screening antibody levels. The seropositive anti-S-RBD IgG ELISA titers represent 3-fold dilutions from 540 to 393,660. Anti-S-RBD IgG dilutional titer and area under the curve (AUC) were quantified. The limit of detection (LOD) was calculated to be half of the lowest AUC for samples with detectable titer (≥1:20) and samples with undetectable titers (1:10) were set be half the LOD.

### Quantification of viral specific anti-RBD and anti-full-length spike in ng/mL

Quantitative antibody measurements were based on an electrochemical immunoassay protocol as previously published(27). A fusion protein of anti-human-IgG coupled to two invertases were used as the electrochemical reporter. Antibody concentrations in ng/mL were obtained by measuring the amount of glucose generated by the protein fusion during immunoassays, based on quantitative dose response-curves built using commercial anti-RBD (Abcam), anti-NTD (ACROBiosystems), and anti-S2 (Novus Biologicals) antibodies.

### SARS-CoV-2 Viral Copy Quantification

Nasopharyngeal specimens obtained at screening were stored in 5 mL of virus transport media at -70°C on site, then shipped to the central storage facility at JHU. RNA was extracted from 200 μL transport media with either the Qiagen viral RNA extraction kit (Qiagen, Hilden, Germany), or the Chemagic Viral DNA/RNA 300 kit 96 (Perkin Elmer, Sheldon CT, USA) followed by real-time reverse transcriptase quantitative PCR (RT-qPCR) assays targeting the SARS-CoV2 nucleocapsid (N) gene and the human RNaseP gene using methods described by the US CDC(28).

### SARS-CoV-2 Virus Neutralization Assay

Plasma neutralizing antibodies were determined against WA-1 (SARS-CoV-2/USA-WA1/2020 EPI_ISL_404895), obtained from BEI Resources, as described previously(29, 30). The limit of viral neutralization detection was at 1:10 titer.

### Statistical analysis

The comparative analysis of anti-S-RBD IgG antibody levels involved calculating the ratio between unique CCP donors and post-transfusion seronegative, unvaccinated recipients. This calculation was performed by dividing the geometric mean (GM) of the area under the curve (AUC) values for donor samples by the corresponding AUC values for the CCP recipients.

We determined correlates of protection based on donor anti-S-RBD IgG levels using two methods: one relying on virus neutralization and the other employing receiver operating characteristic (ROC) analysis. In the first approach, we established a functional cutoff value for binding antibody levels through virus neutralization to distinguish between high and low donor anti-S-RBD IgG AUC levels. It is noteworthy that a virus neutralization antibody at a 1:40 dilutional titer has been previously identified as a correlate of protection in influenza studies(6–8). Initially, we computed the upper limit of the 95% confidence interval for the donor anti-S-RBD IgG AUC geometric mean, corresponding to a donor neutralizing antibody at a 1:40 dilutional titer. The geometric mean was found to be 2,291 AUC, with a lower limit of 1,924 and an upper limit of 2,728 AUC. Considering that the antibody levels of seronegative CCP recipients were approximately 21.3 times lower than those of their respective donors, we extrapolated the functional cutoff point for CCP recipients to be 21.3 times lower than that of donors, resulting in a value of 128 AUC.

Reverse cumulative distribution curves (RCDC) curves were plotted(31) for control and CCP recipients anti-S-RBD post-transfusion ROC analysis. An estimated optimal threshold value from the ROC curve maximizing sensitivity and specificity determined the antibody threshold level for early transfusion. For late transfusions, the maximum percent hospital reduction defined the antibody threshold level.

Spearman correlations were used to evaluate strength of association between titer and AUC units for antibody measurements. The predicted probabilities of hospitalization based on early versus late and high versus low categories were assessed using a Firth’s logistic regression model, chosen due to complete separation in the dataset. Statistical association between hospitalization status and early/high transfusion was assessed by Fisher’s exact test.

Comparisons across groups were performed using Kruskall-Wallis multiple comparisons with Dunn’s post-hoc corrections. We analyzed the antibody kinetics over time among unvaccinated, seronegative recipients using a linear mixed-effects regression model, adjusted for variant, age, sex, and BMI, with anti-S-RBD IgG log_10_(AUC) data. An interaction term was included to examine how antibody levels changed over time by treatment (control or CCP) and hospitalization status. Predicted effects were graphed with 95% confidence intervals. P-values less than 0.05 were considered statistically significant. Analyses were performed using GraphPad Prism 8 (GraphPad Software) or Stata 17 (StataCorp).

## Data Availability

Data is available from individual authors upon request with reply expected in 14 days. Deidentified data from clinical trial has been deposited in the Vivli server for public access.

## Code Availability

Unique software or computational code was not created for this study.

## Author Contributions

HS.P., C.B., A.Yin, J.L., C.C., Y.E., R.F., O.B., J.S., T.G., P.C., J.W., N.N.,M.B., A.T., O.L., S.L.K., D.J.S contributed to the experimental design and procedures for the anti-S-RBD antibody level measurement. K.L., N.M., D.E.F, L.J.A., B.L., D.H., A.C., S.S., E.B., K.G., A.T., O.L., A.P., S.L.K., D.J.S conceived and/or designed the clinical work. M.L., S.Y., I.S., A.J., A.P. contributed to the experimental design and procedure for the viral load and neutralizing antibody measurements.

E.O.G. and N.A.C. performed and supervised, respectively, glucometer-based antibody quantification measurements. A.G.S., G.M.,Y.F., B.F., S.H., A.L., B.M., E.S., S.A., M.H., J.B., J.C., J.H.P., J.G., J.P., P.B, W.R., ME.C, J.H., B.G., V.C., D.C., K.O., M.A., L.H., C.S., D.N.F., M.Z., E.C., J.R., S.G.K., C.M., M.R., A.Yarava, K.L., N.M., A.G., N.K., A.S., D.E.F, D.A.J., L.J.A., D.M.S., B.L., S.E., S.B., T.G., A.Z., D.H., A.C., S.S., E.B., K.G., A.T., O.L., A.P., S.L.K., D.J.S conducted/contributed to the clinical study and/or collected clinical data. HS.P., C.B., A.Yin, J.L., C.C., M.L., S.Y., I.S., A.J., M.R., R.F., O.B., J.S., J.O., Y.B.N., M.D.K.,T.G., P.C., D.H., A.C., K.G., A.T., O.L., A.P., S.L.K., D.J.S contributed to data processing and analyses specific to this work. HS.P., C.B., A.Yin, A.C., A.P., S.L.K., D.J.S drafted the manuscript. All authors provided final approval of the version to be published.

These co-first authors contributed equally: Han-Sol Park, Anna Yin, and Caelan Barranta, with order determined based on role in executing experiments, analyses, and manuscript writing.

## Acknowledgements

Funding

Supported by a contract (W911QY2090012, to Dr. Sullivan) with the Joint Program Executive Office for Chemical, Biological, Radiological and Nuclear Defense of the Department of Defense, in collaboration with the Defense Health Agency; Bloomberg Philanthropies; the State of Maryland; a grant (3R01AI152078-01S1, to Dr. Casadevall) from the National Institutes of Health (NIH) National Institute of Allergy and Infectious Diseases (NIAID); a grant (U24TR001609-S3, to Dr. Hanley) from the NIH National Center for Advancing Translational Sciences; a grant (1K23HL151826NIH, to Dr. Bloch) from the National Heart, Lung, and Blood Institute; the Division of Intramural Research, NIAID, NIH; the Mental Wellness Foundation; the Moriah Fund; Octapharma; the Healthnetwork Foundation; and the Shear Family Foundation. The study sponsors did not contribute to the study design, the collection, analysis, and interpretation of data, and the decision to submit the paper for publication.

## Acknowledgement

We thank all the trial participants who generously gave of their time and donated biologic specimens. We thank Dr. Geeta Sood for a critical review of the manuscript.

## Full methods

### Study design

Our study is a follow-up secondary analysis of a large outpatient, double-blind, randomized clinical trial comparing CCP to control plasma which sought to correlate donor and recipient antibody levels to hospital outcome at 23 centers throughout the United States from June 2020 through September 2021^1^. The trial was halted at 92% (1181/1280) transfusions because of diminishing hospitalizations with increasing vaccinations in fall of 2021. Symptomatic, SARS-CoV-2 test positive, ages 18 or older, regardless of vaccination status or risk factors for severe COVID-19 participants were enrolled within 8 days of symptom onset. The anti-S-RBD IgG dilutional titer and the more precise AUC was quantified in over 5,000 recipient samples at pre-transfusion screening (D-1), 30 minutes post-transfusion (D0), and follow-up visits (D14, D28, D90)^2^. This subgroup analysis was restricted to seronegative, unvaccinated participants. Full study protocol and statistical plan with protocol changes are available with previous publication^1^.

Study sample size was calculated to be 1280 at start of study based on recruitment of an 50% older population with estimate of 30% hospitalization and 15% in age less than 65 years. We assume a one-sided Type I error rate (alpha) of 0.05 as we are interested in superiority and Type II error rate (beta) of 0.2. Therefore, with a sample size of 1344 (1280*1.05 to allow for potential losses) with a target of a minimum ratio of 50:50 for <65:≥65 years of age, we expected to detect at least a 25% reduction in the rate of hospitalization under 80% power and a 30% reduction in rate of hospitalization with 90% power.

### Study Ethics

Johns Hopkins served as the single-IRB (sIRB). For the Center for American Indian Health sites, the protocol was also independently reviewed and approved by the Navajo Nation Health Human Research Review Board and the National Indian Health Service IRB. The protocol was also approved by the Department of Defense (DoD) Human Research Protection Office (HRPO). An independent medical monitor who was unaware of the trial group assignments reviewed all serious adverse events, and an independent panel of three physicians who were unaware of the trial-group assignments adjudicated Covid 19 related hospitalizations and severity. An independent data and safety monitoring board provided interim safety and efficacy reviews.

The trial was conducted in accordance with the principles of the Declaration of Helsinki, the Good Clinical Practice guidelines of the International Council for Harmonisation, and all applicable regulatory requirements. Written and signed informed consent was obtained from all participants.

### Study Population

In this multicenter, double-blind, randomized, controlled trial, we evaluated the efficacy and safety of COVID-19 convalescent plasma, as compared with control plasma, in symptomatic adults (≥18 years of age) who had tested positive for severe acute respiratory syndrome coronavirus 2, regardless of their risk factors for disease progression or vaccination status.

Participants were enrolled within 8 days after symptom onset and received a transfusion within 1 day after randomization. The primary study outcome (reported previously) was COVID-19–related hospitalization within 28 days after transfusion. There were no obvious imbalances between the trial groups in the parent trial with respect to baseline characteristics, including coexisting conditions, COVID-19 vaccination status, vital signs, and clinical laboratory results.

### Study Center(s)

Anne Arundel Medical Center; Ascada Research; Baylor College of Medicine; Johns Hopkins Center for American Indian Health; Johns Hopkins Bloomberg School of Public Health; Johns Hopkins University; Lifespan/Brown University Rhode Island Hospital; Mayo Clinic, Phoenix; MedStar Washington Hospital Center; NorthShore University Health System; The Bliss Group; The Next Practice Group; University of California, Los Angeles Health; University of Alabama at Birmingham; University of California, Irvine Health; University of California, San Diego; University of Cincinnati Medical Center; University of Massachusetts Worcester; University of Miami; University of New Mexico; University of Rochester; University of Texas Health Science Center at Houston; University of Utah Health; Vassar Brothers Medical Center; Wayne State University; Western Connecticut Health Network, Danbury Hospital; Western Connecticut Health Network, Norwalk Hospital.

### Study Donor Plasma

The study qualified donor plasma with SARS-CoV-2 positive antibodies after a 1:320 dilution under FDA IND 19725 protocol. After July 2021, the transfused plasma donor units met the existing FDA Emergency Use Authorization (EUA) criteria for high titer at EUROIMMUN arbitrary unit (AU) over 3.5. Many identical apheresis donor plasma units were transfused into 2, 3, or 4 separate recipients. Plasma from 333 unique CCP donations was transfused into the 592 CCP participants. Seventy-five percent of the donor collections were before September 2020 with more than 90% by January 2021 and the last 25 collections by March 2021. These donor units were previously characterized for full-length anti-Spike IgG geometric mean (GM) titers of 13,053, which corresponded with a more precise area under the curve (AUC) geometric mean of 7938, equaling 243 BAU/mL using the international standards^3^. The median neutralizing antibody (nAb) titer was 80, with a geometric mean titer of 58, and nAb AUC of 51, equaling GM 27 IU/mL^3^. The commercial EUROIMMUN arbitrary units (AU) mean was 6 for the unique donor units^3^.

### Study blinding and allocation

Blinding-Both investigational products—COVID-19 convalescent plasma and control plasma— were matched for ABO compatibility, and the existing labels were covered with labels that read “Thawed plasma (volume), store at 1–6°C; new drug limited by federal (or U.S.) law to investigational use” in order to preserve verification codes.

Allocation-After screening, participants from all 23 sites were randomly assigned in a 1:1 ratio with the use of a central Web-based system and a permuted-block sequence to receive either CCP or control plasma (each administered in a single dose at a volume averaging 214 mL).

Randomization was stratified according to trial site and participant age (<65 years or ≥65 years). The procedures related to randomization of participants at the clinical sites was as follows: Clinical sites collected randomization eligibility and baseline data on the appropriate data collection instruments and entered these data into the database.

The data system confirmed randomization eligibility, issued the next assignment, and relayed treatment assignments to the Data Cordinating Center (DCC) (masked) and blood bank (unmasked).

The data system automatically stored the date and time of assignment, the identity of the clinical site personnel making the assignment, the participant’s ID, and the treatment group assignment.

### Study visits and time periods

In these studies, antibody levels were measured at screen before transfusion, within 30 minutes of transfusion, and various timepoints up to 90 days post-transfusion. Participants were transfused during pre-Alpha (June 3, 2020 to January 31, 2021), Alpha (February 1, 2021 to July 14, 2021), and Delta (July 15 to October 1, 2021) variant periods. There were just three participants transfused from July 2 to July 9, 2021 which decreased the number of false designations. The first Alpha (B1.1.7) confirmed by sequencing was from a participant transfused February 18, 2021.

### Indirect ELISA

The ELISA protocol was adapted from a protocol published by the Florian Krammer laboratory^4^.The 96-well plates (Immulon 4HBX, Thermo Fisher Scientific-Cat#-3855) were coated with anti-S-RBD of the parent strain at a volume of 50 μL of 2 μg/mL diluted antigen in filtered, sterile 1 × PBS (Thermo Fisher Scientific) at 4°C overnight. The coating buffer was removed, and the plates were washed 3 times with 300 μL 1 × PBS plus 0.1% Tween-20 (PBST) wash buffer (Thermo Fisher Scientific) and then blocked with 200 μL PBST with 3% nonfat milk (milk powder, American Bio) by volume for 1 hour at room temperature. All plasma samples were heat-inactivated at 56°C on a heating block for 1 hour before use and diluted 1:2 in PBS. Negative control samples were prepared at 1:10 dilutions in PBST in 1% nonfat milk and plated at a final dilution of 1:100. A mAb against the SARS-CoV-2 Spike protein was used as a positive control (1:5000 dilution; Sino Biological, 40150-D001). Plasma samples were prepared in 3-fold serial dilutions starting at 1:20 in PBST in 1% nonfat milk. Blocking solution was removed, and 100 μL diluted plasma was added in duplicate to the plates and incubated at room temperature for 2 hours. Plates were washed 3 times with PBST wash buffer, and 50 μL of secondary antibody was added to the plates and incubated at room temperature for 1 hour. Antihuman secondary antibody, Fc-specific total IgG HRP (1:5000 dilution; Thermo Fisher Scientific, Invitrogen, A18823), was prepared in PBST plus 1% nonfat milk. Plates were washed, and all residual liquid was removed before the addition of 100 μL SIGMAFAST OPD (o-phenylenediamine dihydrochloride) solution (MilliporeSigma) to each well, followed by incubation in darkness at room temperature for 10 minutes. To stop the reaction, 50 μL 3M HCl (Thermo Fisher Scientific) was added to each well. The OD of each plate was read at 490 nm (OD490) on a SpectraMax i3 ELISA Plate Reader (BioTek Instruments). The positive cutoff value for each plate was calculated by summing the average of the negative values and 3 times the SD of the negatives. Limits of detection (LOD) were set to half the lowest AUC value at or below 20 titer. The anti-S-RBD IgG titer threshold for seronegative was 180 titer or below. The seropositive anti-S-RBD IgG ELISA titers represent 3-fold dilutions from 540 to 393,660.

### Quantification of Viral Specific anti-S-RBD and Anti-Full-Length Spike

The glucose-derived antibody measurements were assessed via an ELISA protocol previously published by the Arroyo laboratory^5^. The protocol was adapted to run on a 96-well plate (Nunc, ThermoScientific 262162). Each well was coated using 50 µL of either S-RBD or full spike protein solutions in PBS, at a concentration of 2.5 and 5.0 ng/mL, respectively. The coating was conducted overnight at 4°C. Wash buffer (WB) was prepared with 1X PBS, pH 7.4 (Fisher Chemical) plus 0.05% Tween (Fisher Bioreagents). Blocking buffer (BB) was prepared by dissolving casein (Fisher Chemical) at 5% w/v in WB. The incubation temperature for each step after coating was 25°C. After coating, the plates were washed three times with WB and then blocked with 200 µL BB for one hour. Then, the plates were washed three times with WB. This procedure was followed by a 30 minute incubation with 50 µL of patient plasma specimens diluted to 1% or 20% with BB, depending on titer levels. Each specimen was interrogated in triplicate. Positive controls (125 and 1,000 ng/mL) and calibration curves (0 to 5,000 ng/mL) for the S-RBD assay employed a commercial mAb against SARS-CoV-2 Spike Glycoprotein S1 (Abcam, ab273073) prepared in 1% or 20% control plasma (to account for both dilutions) diluted in BB. For the full spike protein assay, a 1:1:1 mAb mix against SARS-CoV-2 Spike Glycoprotein S1 (Abcam, ab273073), SARS-CoV-2 Spike S2 (Novus Biologicals, NBP3-07956), and SARS-CoV-2 Spike NTD (ACROBiosystems, SPD-S164) was diluted similarly as for the S-RBD alone. After specimen incubations and washing three times with WB, 50 µL of 0.02 µM of LC15 antibody-invertase fusion protein in BB were added with an incubation time of one hour. The plates were washed three times with WB and once with 1X PBS, pH 7.4. This was followed by a one hour-long incubation with 50µL of 100 mM sucrose (Fisher Chemical) in 1X PBS, pH 5, with glucose concentration measured immediately after using a medical-grade glucometer (Nova Biomedical). Calibration curves were analyzed via nonlinear regression of the Hill isotherm (Igor Pro 8 software) and used to calculate the antibody concentration from the average glucose concentration of each plasma sample.

### SARS-CoV-2 Viral Load

Nasopharyngeal specimens obtained at screen were stored in 5 mL of virus transport media at -70°C on site, then shipped to the central storage facility at Johns Hopkins University. RNA was extracted from 200 mL transport media with either the Qiagen viral RNA extraction kit (Qiagen, Hilden, Germany), or the chemagic Viral RNA/DNA 300 H96 kit with chemagic 360 nucleic acid extraction system (Perkin Elmer), according to manufacturer recommended protocols. Real-time reverse transcriptase quantitative PCR (RT-qPCR) assays targeting the SARS-CoV2 nucleocapsid (N) gene and the human RNaseP gene were performed based on the methods described by the US CDC^6^. Primer and FAM-labelled probe sets for CDC nCoV_N1 and RNaseP assays were purchased from IDT (Integrated DNA Technologies) as part of the SARS-CoV2 Research Use Only RUO qPCR primer and probe kit (part number 10006713, 2019_nCoV RUO kit). Single-plex assays with equivalent volumes of RNA (or Positive Control, Plasmid-RNA Standards or Nuclease Free H2O for No Template Controls (NTCs)) were performed using the TaqPath 1-Step RT-qPCR MasterMix (Applied Biosystems, ThermoFisher Scientific) in a QuantStudio 5 Real-Time PCR system (ThermoFisher Scientific). The SARS-CoV-2 nCoV-N control plasmid comprised the complete nucleocapsid gene of SARS-CoV-2 isolate Wuhan-Hu-1, complete genome (GenBank: NC_045512.2), and the HsRPP30 Positive control contained a portion of the RNAseP (RPP30) gene. Both plasmid controls were purchased from IDT. Standards for quantitative analysis were prepared from serial dilutions of the nCoV-N and HsRPP30 plasmid controls for which target copy number was known. The range covered was 200,000 copies to 320 copies. Standard curve analysis of nCoV_N1 Ct values was performed by the QuantStudio Design and Analysis software to determine RNA copies of viral genome. Only samples with quantities within the standard curve range were given a COVID-19 call/score “positive”. A Ct value for the RNaseP gene was used to verify that human RNA was present in each specimen. For samples that did not amplify viral genome or any host cell RNA, a repeat RT-qPCR was performed and subsequently assigned as “undetermined”.

### SARS-CoV-2 Virus Neutralization Assay

Plasma neutralizing antibodies were determined against WA-1 (SARS-CoV-2/USA-WA1/2020 EPI_ISL_404895), which was obtained from BEI Resources, as described previously^7,8^. Two-fold dilutions of plasma (starting at a 1:20 dilution) were made and infectious virus was added to the plasma dilutions at a final concentration of 1 × 10^5^ TCID_50_/mL (100 TCID_50_ per 100 μL). The samples were incubated with the virus for 1 hour at room temperature, and then 100 μL of each dilution was added to 1 well of a 96-well plate of VeroE6-TMPRSS2 cells in hexaplicate.

The cells were incubated for 6 hours at 37°C, 5% CO_2_. The inocula were removed, fresh infection media (IM) was added, and the cells were incubated at 37°C, 5% CO_2_ for 2 days. The cells were fixed by the addition of 100 μL of 4% formaldehyde per well, incubated for at least 4 hours at room temperature, and then stained with Napthol Blue Black (MilliporeSigma). The neutralizing antibodies titer was calculated as the highest serum dilution that eliminated the cytopathic effect in 50% of the wells (NT50), and the AUC was calculated using Graphpad Prism.

### Primary Study Statistical Analysis

The statistical analysis plan, included with the trial protocol at NEJM.org, was finalized before database lock and unblinding. We calculated the risk difference and the restricted mean survival time (the expected mean time to hospitalization or death by 28 days) in a modified intention-to-treat analysis that excluded participants who did not receive transfusion of convalescent plasma or control plasma. We estimated the cumulative incidence using the doubly robust estimator based on a targeted minimum loss–based estimator. In order to increase the precision of estimates and to account for potential dependent censoring, the analyses were adjusted for baseline variables that were potentially related to the primary outcome. In order to determine which prespecified candidate variables to include, we conducted variable selection using the random survival forest method in the entire sample while we were unaware of the trial-group assignments. We used imputation for missing values in an algorithm to select covariates for inclusion in a targeted minimum loss–based estimation model. A time-to-event analysis was based on the period from the time of transfusion until an outcome occurred. A two-sided test with a type I error of 0.05 was used to determine statistical significance.

### Secondary Analysis Statistical analysis

The ratio of anti-S-RBD IgG antibody levels between unique CCP donors and D0 seronegative, unvaccinated recipients was calculated by dividing the geometric mean (GM) of donor AUC values by that of the CCP recipients.

We established correlates of protection using donor anti-S-RBD IgG levels by to methods—one based in virus neutralization and another using ROC analysis. For the first method, we established a functional cutoff value for binding antibody levels based in virus neutralization to delineate between high and low donor anti-S-RBD IgG AUC levels. Virus neutralization antibody at 1:40 dilutional titer has previously been reported as a correlate of protection in previous influenza studies [insert citation]. First, we calculated the upper limit of 95% confidence interval for the donor anti-S-RBD IgG AUC geometric mean at which the donor nAb is at 1:40 dilutional titer. We identified a GM of 2291 AUC with a lower limit of 1924 and upper limit of 2728 AUC. Recognizing that the antibody levels of seronegative CCP recipients were approximately 21.3 times lower than their respective donors, we further inferred the functional cutoff point for CCP recipients to also be 21.3 times lower than that of donors (128 AUC).

The RCDC curves were plotted^9^ for control and CCP recipients anti-S-RBD post-transfusion. To calculate the antibody threshold level for early transfusion, a logistic regression model with hospitalization as the outcome and post-transfusion antibody level as the predictor was fitted for seronegative and unvaccinated participants who received early treatment. A ROC curve was plotted using the logistic regression model. An estimated optimal threshold value from the ROC curve that maximizes sensitivity and specificity was established by Youden’s J statistics. The optimal antibody level associated with the estimated optimal threshold value from ROC was determined using the fitted logistic regression model. For late transfusions, the maximum percent hospital reduction on the two curves determined the antibody threshold level.

Spearman correlations were used to evaluate strength of association between titer and AUC units for antibody measurements. Predicted probabilities of hospitalization by the early versus late and high versus low categories were assessed using a Firth’s logistic regression model due to complete separation in the dataset. Longitudinal seronegative recipient antibody data were first log_10_-transformed and analyzed using a linear mixed-effects regression model, adjusted for variant, age, sex, and BMI. An interaction term was included to examine how antibody levels changed over time by treatment (control or CCP) and hospitalization status. Predicted effects were graphed with 95% confidence intervals. Statistical analyses were performed using GraphPad Prism 8 (GraphPad Software) and Stata 17 (StataCorp).

---

**Supplemental Figure 1.**
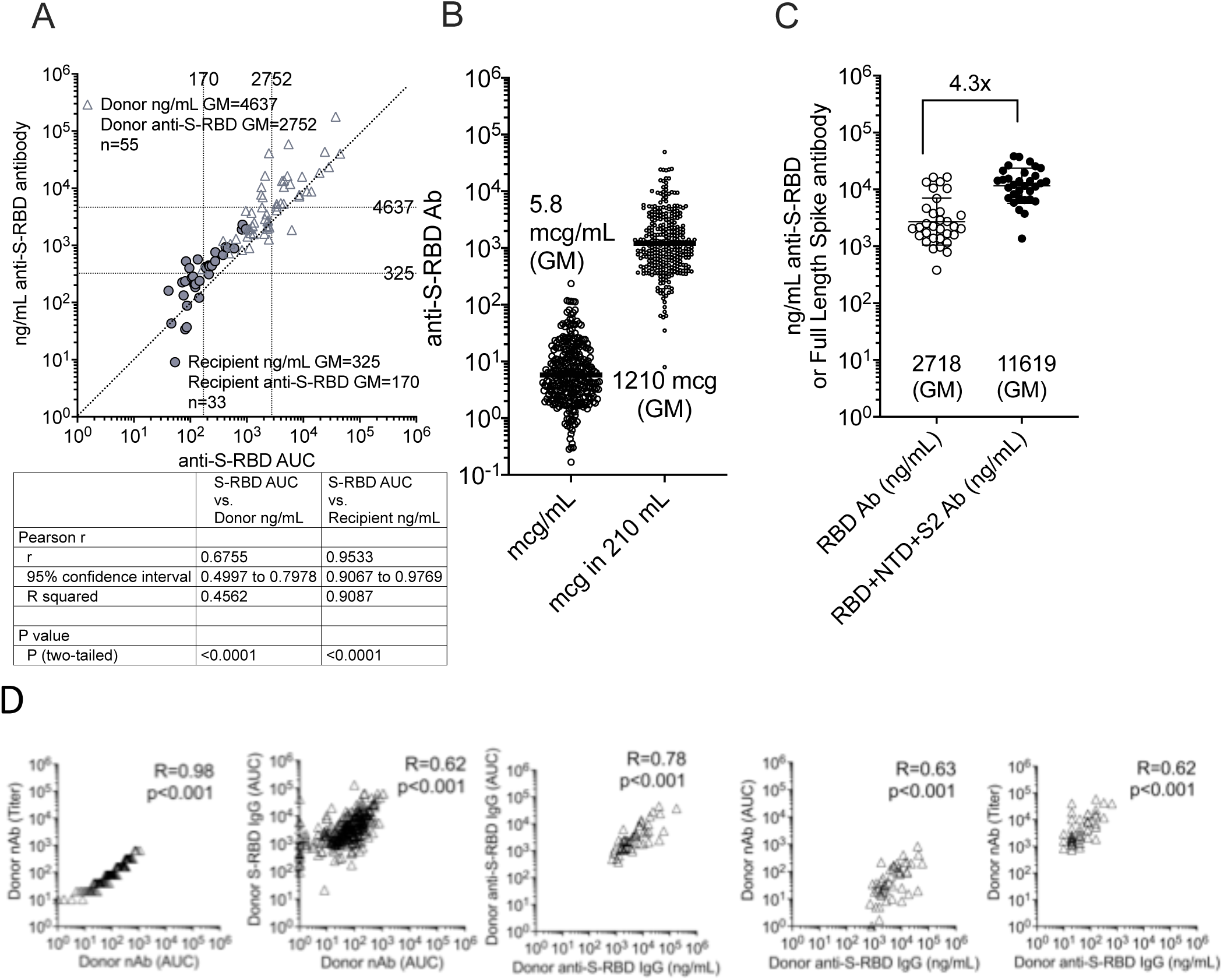
Conversion of anti-S-RBD AUC to ng/mL viral specific antibody. A) Determination of ng/mL of anti-S-RBD specific antibody levels in both post-transfusion recipients (n=33) and donors (n=55) with correlation to anti-S-RBD AUC. The ng/mL units were 1.8 fold greater than anti-S-RBD AUC comparing geomeans of the combined 88 samples tested. B) Using the strong correlation of determined ng/mL to S-RBD AUC, the 319 anti-S-RBDs AUC of unique donor units were converted to ng/mL (RBD AUCx1.8= ng/mL S-RBD antibody) and multiplied by 210 the average volume of transfusion to approximate a 1210 geomean for total mcg of viral specific S-RBD. C) Total full length spike ng/mL is approximately 4.3 times S-BD ng/mL (n=31) which translates to 5.2 total mg spike viral specific antibody dose per donor unit. D) Spearman correlation of donor anti-S-RBD IgG across different units of measurement (e.g., ng/mL, AUC, titer).

**Supplemental Figure 2.**
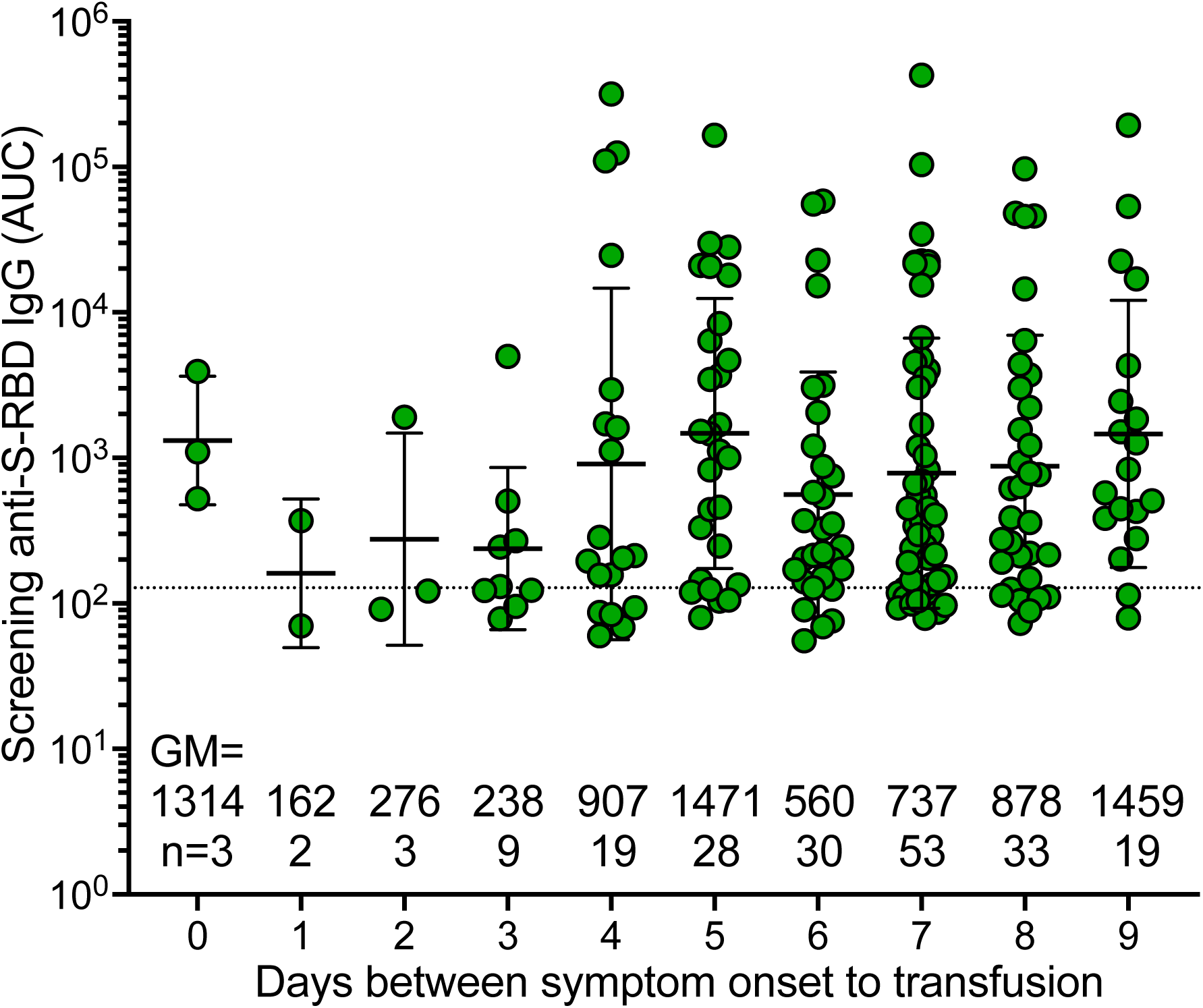
Screen seropositive participants antibody levels stratified by transfusion days from symptom onset. The 199 unvaccinated seropositive participant screen pretransfusion antibody levels stratified by days from symptom onset to transfusion. All point estimates are shown with error bars indicating the geometric mean with geometric SD. Numbers above the x-axis represent geometric mean (GM), the number in the group (n). The dashed line in B-E represents the upper post-transfusion 128 AUC recipient’s threshold. All point estimates are shown with error bars indicating the mean with SD.

**Supplemental Figure 3.**
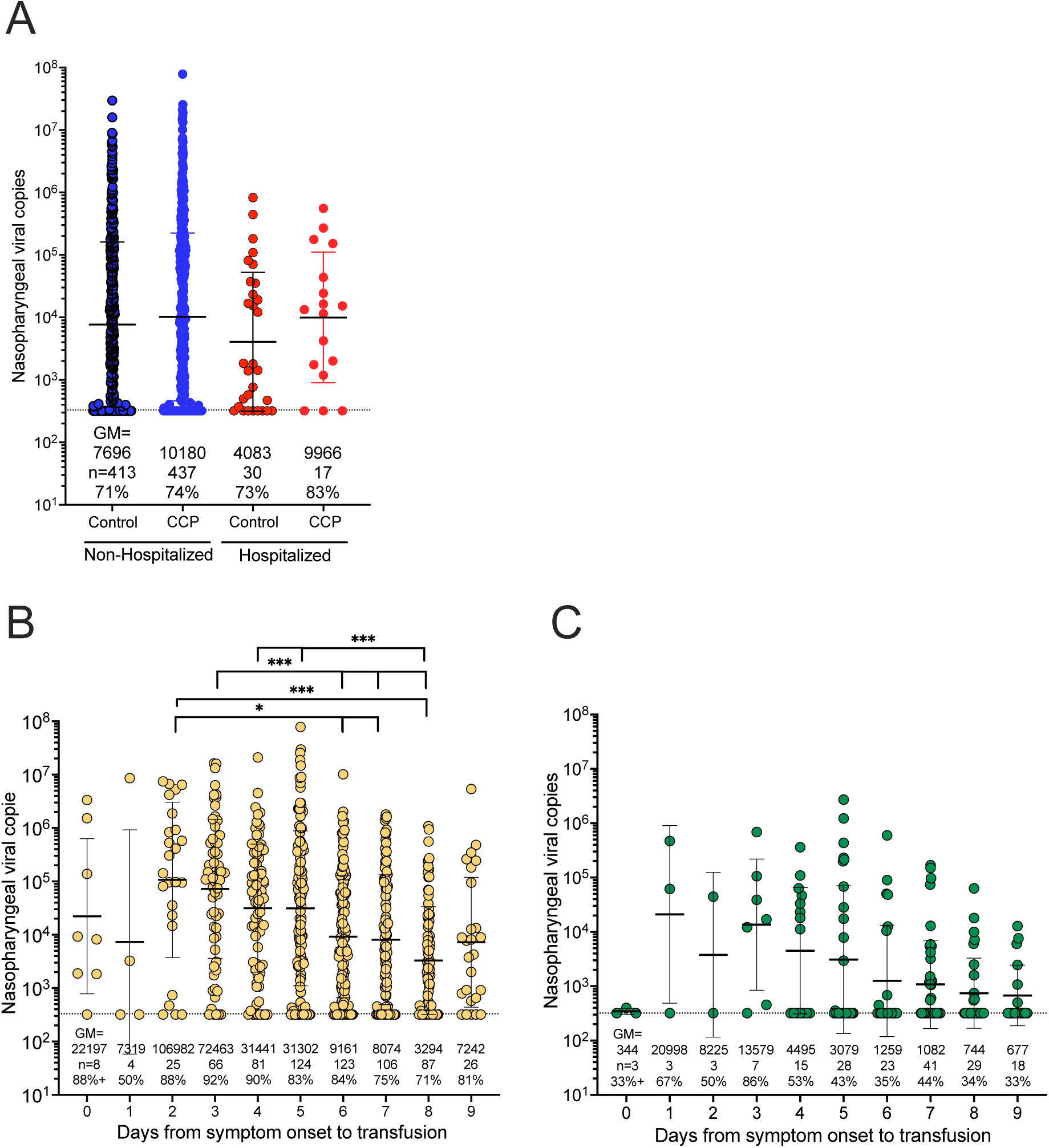
Screen pre-transfusion nasal swab viral load determinations. segregated by A) CCP or control plasma administration in those hospitalized or not hospitalized. Unvaccinated pre-Delta period participants were segregated into B) seronegative and C) seropositive populations by symptom duration in days to transfusion. Numbers above the x-axis represent geometric mean (GM), the number in the group (n), and percentage of PCR-positive samples (%) for each category. *** p<0.001, ** p=0.002 and * p=0.033 by non-parametric Kruskal-Wallis multiple comparisons test with Dunn’s post-hoc corrections. All point estimates are shown with error bars indicating the GM with SD. The dashed lines indicate samples below the limit of detection of 330 viral copies.

**Supplemental Figure 4.**
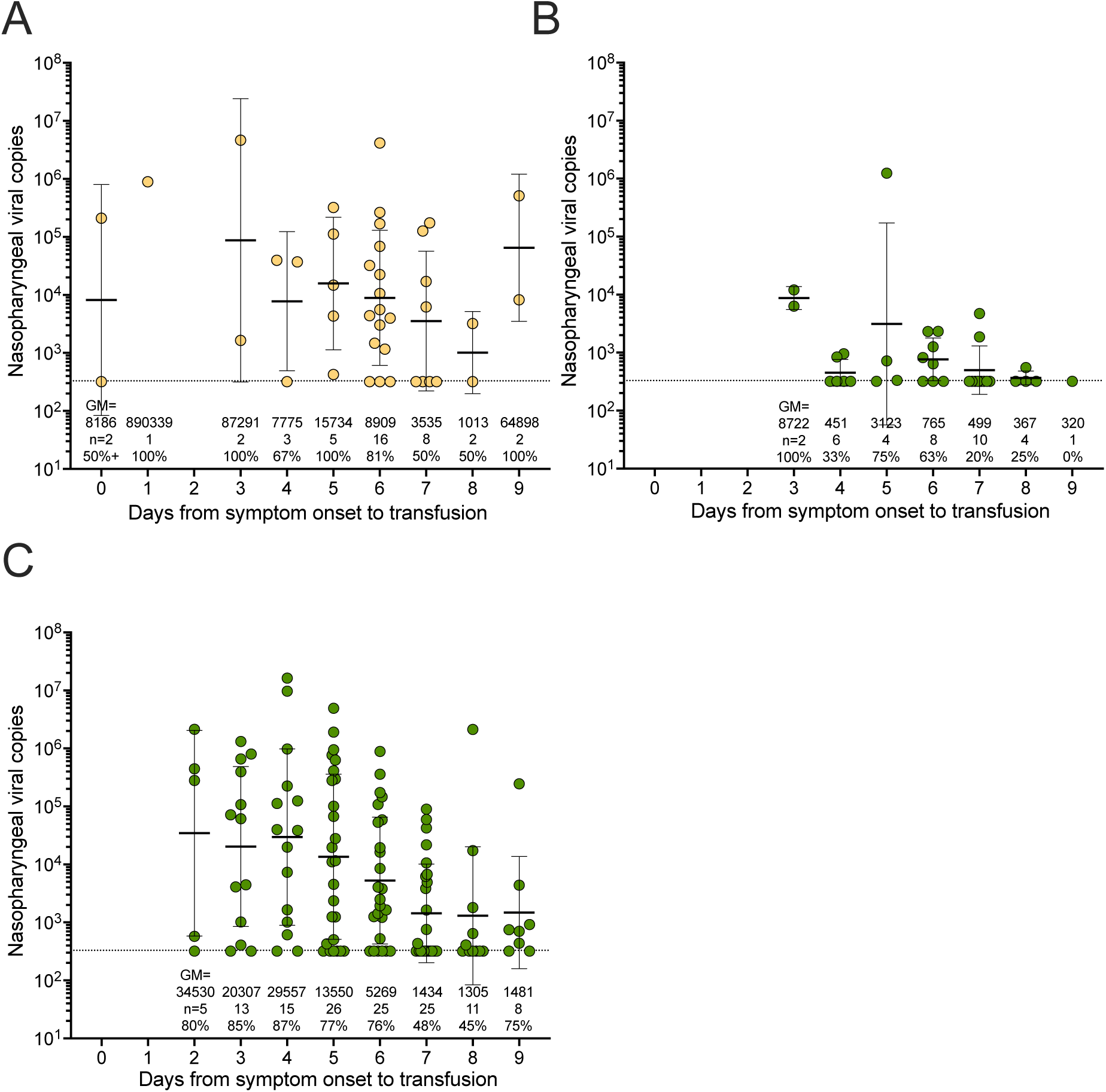
Screen viral loads during the Delta period. During the Delta period there were only 77 (34%) participants unvaccinated to segregate into A) seronegative (n=42) and B) seropositive (n=35) groups by duration from symptom onset to transfusion. C) During the Delta period fully vaccinated participants (n=128) were antibody positive with an additional single recipient fully vaccinated, but seronegative with nasal viral load on day 0 of 320 (not graphed). All point estimates are shown with error bars indicating the GM with SD. Numbers above the x-axis represent geometric means (GM), the number in the group (n), and the percentage of samples PCR positive (%). The dashed lines indicate samples below the limit of detection of 330 viral copies.

**Supplemental Figure 5.**
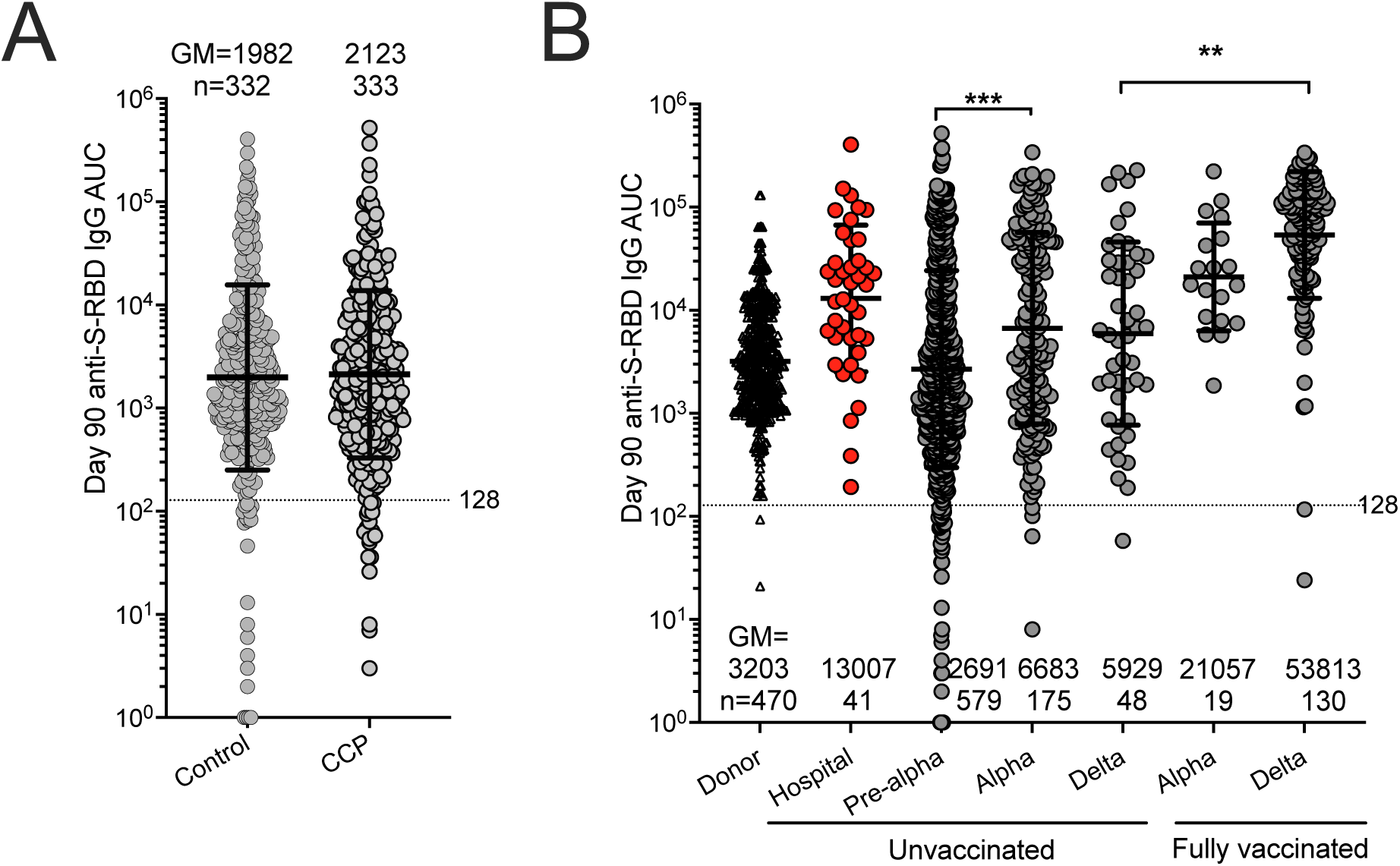
Antibody levels three months post-transfusion. Unvaccinated recipients anti-S-RBD AUC antibody levels at Day 90 post-transfusion (excluding the 165 vaccinated during the follow-up visits) separated by A) CCP and control recipients B) both CCP and control recipients by SARS-CoV-2 variant period and vaccination status. Clear squares indicate donor, red squares indicate hospitalized recipients, and gray squares indicate both CCP and control non-hospitalized recipients. All point estimates are shown with error bars indicating the GM with SD. Numbers above the x-axis represent each category’s geometric mean (GM) and number in the group (n). The dashed line in A, B represents the upper portion post-transfusion 150 AUC recipient’s threshold, GM donor 3286 AUC and GM donor 6678 titer. *** p<0.001, by non-parametric Kruskal-Wallis multiple comparisons test with Dunn’s post-hoc corrections.

**Supplemental Table 1.**
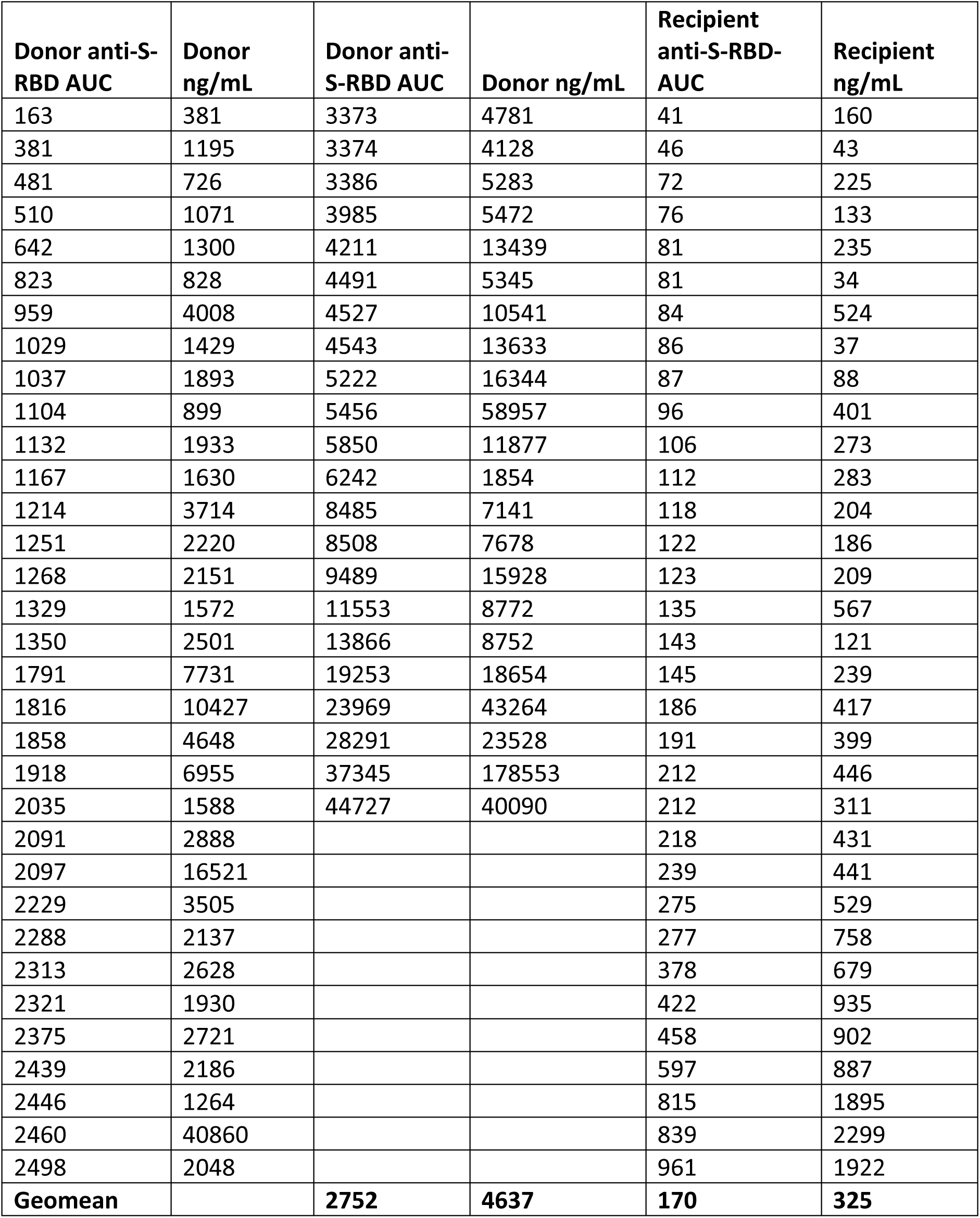
Data for ng/mL glucose based determination of viral specific antibodies.

| **Donor anti-S-RBD AUC** | **Donor ng/mL** | **Donor anti-S-RBD AUC** | **Donor ng/mL** | **Recipient anti-S-RBD-AUC** | **Recipient ng/mL** |
| --- | --- | --- | --- | --- | --- |
| 163 | 381 | 3373 | 4781 | 41 | 160 |
| 381 | 1195 | 3374 | 4128 | 46 | 43 |
| 481 | 726 | 3386 | 5283 | 72 | 225 |
| 510 | 1071 | 3985 | 5472 | 76 | 133 |
| 642 | 1300 | 4211 | 13439 | 81 | 235 |
| 823 | 828 | 4491 | 5345 | 81 | 34 |
| 959 | 4008 | 4527 | 10541 | 84 | 524 |
| 1029 | 1429 | 4543 | 13633 | 86 | 37 |
| 1037 | 1893 | 5222 | 16344 | 87 | 88 |
| 1104 | 899 | 5456 | 58957 | 96 | 401 |
| 1132 | 1933 | 5850 | 11877 | 106 | 273 |
| 1167 | 1630 | 6242 | 1854 | 112 | 283 |
| 1214 | 3714 | 8485 | 7141 | 118 | 204 |
| 1251 | 2220 | 8508 | 7678 | 122 | 186 |
| 1268 | 2151 | 9489 | 15928 | 123 | 209 |
| 1329 | 1572 | 11553 | 8772 | 135 | 567 |
| 1350 | 2501 | 13866 | 8752 | 143 | 121 |
| 1791 | 7731 | 19253 | 18654 | 145 | 239 |
| 1816 | 10427 | 23969 | 43264 | 186 | 417 |
| 1858 | 4648 | 28291 | 23528 | 191 | 399 |
| 1918 | 6955 | 37345 | 178553 | 212 | 446 |
| 2035 | 1588 | 44727 | 40090 | 212 | 311 |
| 2091 | 2888 |  |  | 218 | 431 |
| 2097 | 16521 |  |  | 239 | 441 |
| 2229 | 3505 |  |  | 275 | 529 |
| 2288 | 2137 |  |  | 277 | 758 |
| 2313 | 2628 |  |  | 378 | 679 |
| 2321 | 1930 |  |  | 422 | 935 |
| 2375 | 2721 |  |  | 458 | 902 |
| 2439 | 2186 |  |  | 597 | 887 |
| 2446 | 1264 |  |  | 815 | 1895 |
| 2460 | 40860 |  |  | 839 | 2299 |
| 2498 | 2048 |  |  | 961 | 1922 |
| **Geomean** |  | **2752** | **4637** | **170** | **325** |

**Supplemental Table 2.** Firth’s Logistic Regression Contrasts and P-values.

| Method of Cutoff | Quadrant Category | Predicted Probability | P-value |
| --- | --- | --- | --- |
| Virus Neutralization<br>Antibody Cutoff | Early/Low | 0.0636567 | 0.008 |
|  | Early/High | 0.0086185 | 0.465 |
|  | Late/Low | 0.0457482 | 0.045 |
|  | Late/High | 0.0605447 | 0.007 |
| ROC Cutoff | Early/Low | 0.0673155 | 0.008 |
|  | Early/High | 0.0082876 | 0.462 |
|  | Late/Low | 0.0567985 | 0.002 |
|  | Late/High | 0.0446232 | 0.174 |

| Method of Cutoff | Quadrant Comparisons | P-value |
| --- | --- | --- |
| Virus Neutralization<br>Antibody Cutoff | Early/Low vs Early/High | 0.145 |
|  | Late/Low vs Early/High | 0.237 |
|  | Late/High vs Early/High | 0.160 |
| ROC Cutoff | Early/Low vs Early/High | 0.124 |
|  | Late/Low vs Early/High | 0.153 |
|  | Late/High vs Early/High | 0.276 |

**Supplemental Table 3.** Fishers Exact table results for unvaccinated seronegative participants.

| Unvaccinated seronegative | hospital | no hospital | total | Hospital percent | p to all seroneg control | row p to early cont | row p to late control | p to all other CCP | p to early low CCP |
| --- | --- | --- | --- | --- | --- | --- | --- | --- | --- |
| all controls | 31 | 238 | 268 | 12.0 |  |  |  |  |  |
| early control | 20 | 147 | 167 | 12.0 |  |  |  |  |  |
| late control | 11 | 191 | 201 | 5.0 |  |  |  |  |  |
| donor early high | 1 | 87 | 88 | 1.1 | **0.002** | **0.002** |  | 0.13 | 0.21 |
| donor early low | 4 | 82 | 86 | 4.7 | 0.09 | 0.07 |  |  |  |
| donor late high | 6 | 98 | 104 | 5.8 | 0.12 |  | 0.78 |  |  |
| donor late low | 4 | 79 | 83 | 4.8 | 0.09 |  | 1 |  |  |
| donor early low and late | 14 | 259 | 273 | 5.1 |  |  |  |  |  |
| recip nAb early high | 0 | 85 | 85 | 0.0 | **0.0002** | **0.0003** |  | **0.03** | 0.058 |
| recip nAb early low | 5 | 81 | 86 | 5.8 | 0.21 | 0.18 |  |  |  |
| recip nAb late high | 7 | 107 | 114 | 6.1 | 0.18 |  | 0.8 |  |  |
| recip nAb late low | 3 | 73 | 76 | 3.9 | 0.08 |  | 1 |  |  |
| recip nAb early low and Late (high/low) | 15 | 261 | 276 | 5.4 |  |  |  |  |  |
| recip rcdc early high | 0 | 94 | 94 | 0.0 | **0.0001** | **0.0001** |  | **0.01** | **0.017** |
| recip rcdcearly low | 5 | 72 | 77 | 6.5 | 0.3 | 0.256 |  |  |  |
| recip rcdc late high | 1 | 39 | 40 | 2.5 | 0.1 |  | 0.69 |  |  |
| recip rcdc late low | 9 | 141 | 150 | 6.0 | 0.11 |  | 0.81 |  |  |
| recip rcdc early low and Late (high/low) | 15 | 252 | 267 | 5.6 |  |  |  |  |  |

